# A hybrid computer vision model to predict lung cancer in diverse populations

**DOI:** 10.1101/2024.10.07.24315011

**Authors:** Abdul J. Zakkar, Nazia Perwaiz, Vikram Harikrishnan, Weiheng Zhong, Vijeth Narra, Alex Krule, Farah Yousef, Daniel Kim, Mason Burrage-Burton, Abdul Afeez Lawal, Vijayakrishna K. Gadi, Mark C. Korpics, Sage J. Kim, Zhengjia Chen, Aly A. Khan, Yamilé Molina, Yang Dai, G. Elisabeta Marai, Hadi Meidani, Ryan H. Nguyen, Ameen A. Salahudeen

## Abstract

**PURPOSE:** Disparities of lung cancer incidence exist in Black populations and screening criteria underserve Black populations due to disparately elevated risk in the screening eligible population. Prediction models that integrate clinical and imaging-based features to individualize lung cancer risk is a potential means to mitigate these disparities.

**PATIENTS AND METHODS:** This Multicenter (NLST) and catchment population based (UIH, urban and suburban Cook County) cross-sectional study utilized participants at risk of lung cancer with available lung CT imaging and follow up between the years 2015 and 2024. 53,452 in NLST and 11,654 in UIH were included based on age and tobacco use based risk factors for lung cancer. Cohorts were used for training and testing of deep and machine learning models using clinical features alone or combined with CT image features (hybrid computer vision).

**RESULTS:** An optimized 7 clinical feature model achieved ROC-AUC values ranging 0.64-0.67 in NLST and 0.60-0.65 in UIH cohorts across multiple years. Incorporation of imaging features to form a hybrid computer vision model significantly improved ROC-AUC values to 0.78-0.91 in NLST but deteriorated in UIH with ROC-AUC values of 0.68-0.80, attributable to Black participants where ROC-AUC values ranged from 0.63-0.72 across multiple years. Retraining the hybrid computer vision model by incorporating Black and other participants from the UIH cohort improved performance with ROC-AUC values of 0.70-0.87 in a held out UIH test set.

**CONCLUSION:** Hybrid computer vision predicted risk with improved accuracy compared to clinical risk models alone. However, potential biases in image training data reduced model generalizability in Black participants. Performance was improved upon retraining with a subset of the UIH cohort, suggesting that inclusive training and validation datasets can minimize racial disparities. Future studies incorporating vision models trained on representative data sets may demonstrate improved health equity upon clinical use.

## INTRODUCTION

Lung cancer treatment in the United States carries health disparities in minority populations and are especially pronounced in Black individuals^1^. Low-dose CT (LDCT) lung cancer screening shifts diagnosis to early-stage disease and is associated with reduced mortality^2,3^, but stringent eligibility criteria from the United States Preventive Services Task Force (USPSTF)^4^ exacerbates disparities in Black populations^4–6^. These criteria, specifically regarding age and duration of tobacco usage, were derived from risk/benefit analysis of the ∼ 92% White National Lung Screening Trial (NLST) cohort. This results in exclusion of at-risk Black patients from screening. The eligibility to incidence ratio measures this disparity, and according to current USPSTF guidelines, White populations often have >100 eligible patients screened for one incidence of lung cancer, whereas in Black populations this is reduced to approximately 50 eligible patients for every diagnosed case of lung cancer^4^.

Our institution^7^ and others^6,8,9^ have identified that individualized selection using clinical feature-based risk models improved lung cancer screening eligibility and sensitivity in minoritized populations as compared to USPSTF guidelines. Moreover, individualized lung cancer risk prediction from LDCT images has been achieved using Convolutional Neural Networks (CNNs)^10–13^. Taken together, risk prediction models have the potential to guide screening selection and eligibility, aide the ordering physician and radiologist in determining follow up imaging interval and mitigate lung cancer disparities in early detection and survival. However, these models have been trained and evaluated on cohorts which underrepresent minorities. To advance these risk models for implementation, their generalizability requires assessments in minority populations including Black patients who have the highest annual lung cancer incidence in the United States^14^.

In this study, we developed a hybrid computer vision model that integrates clinical and imaging features to predict individual lung cancer risk. We evaluated the hybrid computer vision model’s predictions in both the National Lung Screening Trial (NLST) cohort and assembled a diverse urban and suburban real-world patient population across multiple sites in Cook County within the Chicago metropolitan area (University of Illinois Health, UIH) altogether representing 65,106 participants including 8,823 self identified as Black. Model risk predictions were evaluated against ground-truth lung cancer diagnoses to determine whether performance was accurate and equitable across various subpopulations.

## METHODS

Research analysis was carried out under approved UIC IRB protocols 2023-1321 and 2023-1377. The NLST is a multicenter cohort and has been previously described^2^. Cook county is racially, socioeconomically, and geographically diverse and has regions with higher rates of lung cancer, predominantly in areas populated by Black patients^15–17^. We assembled a real-world cohort of 11,654 participants treated in urban and suburban settings in the city of Chicago as well as greater Cook County. This population based cohort, including 6,447 Black patients with history of smoking and/or lung cancer, was filtered based on available lung/chest CT studies.

### Collecting Data: NLST

Deidentified NLST data was accessed from the National Cancer Institute Cancer Data Access System approved under approved UIC IRB Protocol 2023-1321.

### Collecting Data: UIH Lung Cancer Screening

To assess the hybrid model in the UIH population, we assembled a retrospective cohort of patients with a history of smoking under approved UIC IRB protocol 2023-1377. Inclusion criteria for this cohort is listed in **eTable 1**. Briefly, participants were placed into the cohort to mimic USPSTF lung cancer screening eligibility criteria with ages 50-80 and a history of tobacco use, and/or based on ICD codes for smoking cessation counseling and cigarette smoking, and through documented history of smoking. ICD-9 code 305.1 and ICD-10 code F17 were used to include LDCT procedures to capture participants who may have had missing data in tobacco data tables from the EHR. LDCT associated DICOMS were chosen through a filtering schema (**eFigure 1**). A diagnosis of lung cancer was determined based on ICD codes of neoplasm of bronchus or lung (ICD-9 162, ICD-10 C34), since 10-15 percent of patients at UIH receive stereotactic body radiation therapy without biopsy proven lung cancer. Deidentified data from positive cases were verified by A.A.S., a board-certified medical oncologist with sub specialization in thoracic oncology. Those without a diagnosis of lung cancer and engagement in the EHR during the retrospective window were assumed to have no lung cancer at the time of the preparation of this manuscript. Median duration of follow-up between LDCT scans was 431 days and the median number of LDCT scans per participant with at least one LDCT was 1 (the median is 5 if each convolution kernel and reconstruction of a given LDCT exam is counted separately). Samples belonging to patients who died or were lost to follow-up before a given time point were omitted. For example, if the same patient died or was lost to follow-up 4.5 years after a CT scan was performed, then the ground truth set for CT sample would be [0, 1, 1, 1, NA, NA].

### Generating Ground Truth

The ground truth is represented by six values (1 or 0) indicating the presence or absence of a lung cancer diagnosis 1 to 6 years after a CT scan was performed. Using the Python programming language, a data frame was created which represents the ground truth associated with each LDCT in both the NLST and UIH data sets. The time of each CT event was compared to the time of the lung cancer diagnosis event of the associated participant. If the difference is less than or equal to a given number of years, it is labeled as a positive sample (1), otherwise it is labeled as a negative sample (0). If the difference is less than or equal to a given number of years, it is labeled as a positive sample (1), otherwise it is labeled as a negative sample (0). For example, if a CT scan was performed 500 days before lung cancer diagnosis, this means it occurred with 2, 3, 4, 5, and 6 years of diagnosis, but not within 1 year. Therefore, the ground truth set for this CT sample would be [0, 1, 1, 1, 1, 1]. However, if the sample has no known lung cancer diagnosis event, and the number of years exceeds the number of days to last follow-up event (or in the case of UIH data, the date 3/16/2024), then the sample is excluded in that specific follow up period due to a lack of information in determining the ground truth. Over years 1-6, 1888, 3128, 3828, 4056, 4476, and 4884 studies were removed from the UIH cohort respectively (i.e. 5,980 LDCT studies initially, then 4092 with one year of follow up, etc.). The NLST data set includes the number of days to the last known cancer status of each participant, and this was used in a similar manner to remove 4, 35, 159, 280, 461, and 1082 samples (out of 11,980) from the NLST data test set for each respective year.

### Training and Test set selection in NLST

A training set from the NLST was created using the same split as previously described in Ardila et al.^10^ for the NLST dataset. Specifically, participants included in the Ardila et al. test set (n = 2,328) were allocated to our test set and were kept entirely unseen during training and model development. The remaining participants were randomly assigned to either the training set or the development set with the ratio of 80-20, the assignment is performed at the participant level to avoid data leakage between sets. Each participant contributes one or more LDCT scans, with each sample representing an individual patient. In addition, data from participants in the chest ray arm of NLST were also incorporated where appropriate for auxiliary comparisons, such as the evaluation of the PLCOm2012 risk model, which relies solely on clinical features and not imaging. The SVM models which incorporate imaging features via ResNet scores were also trained using the same training set, however each sample represents a single CT scan rather than an individual. The specific test set IDs will be made available in a Zenodo repository upon publication of this manuscript.

### Training and Test set selection in UIH

Data from the UIH cohort consisted of various Digital Imaging and Communications in Medicine (DICOM) types, a digital format for storing CT scans and associated metadata. LDCTs were selected using a schema detailed in **eFigure 1**. Coded UIH subject IDs for the training and test split for the Hybrid model will be made available in a University of Illinois Indigo Repository upon publication of this manuscript.

### Prediction with PLCO_m2012_

PLCO_m2012_ is a logistic regression model for lung cancer risk inference and was evaluated on NLST data. The PLCO_m2012_ model was published in 2012, and was created using a set of 11 chosen features to predict the probability of lung cancer diagnosis for a given participant within 6 years: age, race, education, Body-Mass Index (BMI), presence of Chronic Obstructive Pulmonary Disease (COPD), personal history of cancer, family history of lung cancer, smoking status, cigarettes per day, duration of smoking, and duration of quitting. All 11 features are directly represented or can be extracted from the data in the NLST. The PLCO_m2012_ logistic regression model is represented in an R package created by the authors of PLCO_m2012_^16^. This was translated to Python code to facilitate incorporation with the remainder of the code base. When using PLCO_m2012_ for prediction, each sample represents an individual participant. The model was used to generate predictions for probability of lung cancer diagnosis within 6 years only (it was not used to evaluate performance within 1 to 5 years) as intended by its authors^18^.

### SVM Prediction

Clinical data elements from the NLST were evaluated for inclusion in a SVM using the Scikit-Learn Python library^19^. 11 features were retained after recursive feature elimination using linear kernel. We started with a large set of initial features from those available in NLST, as well as some generated features which were calculated from existing ones, such as BMI. Recursive feature elimination showed best results when using 11-13 features. Six unique SVMs (one per year) were then trained to make predictions 1-6 years post CT scan, with each sample being an individual participant. Each of the 6 SVM models was then evaluated on the test sets using the corresponding ground truths. Additional SVM models were created using a truncated set of features for more practical utility using highly populated EHR fields, and to also allow for comparison between UIH and NLST cohorts where features had a high preponderance data missingness. The training and prediction pipeline of SVM method is depicted in **eFigure 2**.

### XGBoost Survival (Cox) Model Implementation

A Cox Proportional Hazards survival model was implemented using XGBoost, trained on 63 clinical variables to predict patient risk over time in the NLST training data set. The XGBoost model was configured to optimize the Cox partial likelihood, specifically designed for censored survival data, and outputs a risk score for each patient that represents the log of the hazard (log hazard ratio). After training, the predicted log hazard ratios from the XGBoost model were not used directly as absolute risk probabilities since they were unbounded, but were instead used as single covariates in a secondary Cox proportional hazards model, which was fit using the Lifelines library. This second step enables the estimation of baseline survival function and allowed translation of the XGBoost-derived risk scores into individualized, time-specific survival probabilities for each participant. Specifically, the probability of experiencing the event (e.g., cancer diagnosis) was estimated within 1 to 6 years by calculating the survival probability at each yearly time point and then converting it to a risk probability (risk = 1 - survival probability).

### XGBoost, Decision Tree, & Transformer models

XGBoost, Decision Tree, and Transformer models from the Scikit-Learn Python library^19^ were trained were trained with 63 clinical features on the NLST clinical training dataset using default parameters and tested on NLST test dataset as above.

### ResNet Prediction

DICOM data files were evaluated by a ResNet-18 CNN model, trained on the ImageNet database with parameters locked and further trained upon the NLST cohort as previously described^10^. The ResNet model, Sybil, was run with default parameters on the same subset of the NLST cohort comprised of 11,980 LDCT studies from 2,328 participants. The resulting output from each LDCT image was a set of 6 values indicating the probability that lung cancer was diagnosed within 1 to 6 years from the date of the CT study. In the UIH cohort, the ResNet model was used on 4,092 LDCT studies from 1,104 participants. Each prediction score was then compared with the ground truth values to determine the model’s performance, broken down into 6 performance evaluations by years 1 to 6.

### SVM + ResNet Hybrid Model Prediction

For the hybrid computer vision model, SVM learning was used to train six unique models for years 1-6 post imaging including the corresponding ResNet inferred risk for that given year. Each ResNet + SVM model incorporates the ResNet risk inference of the current year and that the training and testing samples are individual DICOMs associated with the clinical features of each subject ID at baseline study entry. The training and test sets used the same allocation of NLST participants used to train the previous ResNet model, and a 70:30 split of participants for the UIH training cohort. To ensure equal comparison between the various models, training and test sets had identical allocation of participants and corresponding DICOM files.

### Cross Layer Attention Model Predictions

In addition to the late fusion approach with the SVM + ResNet Hybrid Model, we also evaluated multiple approaches for integrating clinical and imaging data to predict the risk of lung cancer. We investigated early fusion strategies that were enabled using cross-attention architectures that integrated clinical variables with 512-dimensional LDCT embeddings. Three distinct cross-attention architectures were explored:

1. Graph Neural Network (GNN)-based cross-attention model, a graph convolutional network (GCN) model, utilizing late fusion approach for interactions between modalities, with cross-attention layers enabling feature exchange between clinical and imaging representations.
2. Custom cross-attention model, a two-layer cross-attention architecture explicitly modeled pairwise interactions between clinical and imaging features without graph structure. The design allowed selective feature weighting across modalities but offered less structural prior than the GNN-based method.
3. Transformer-based cross-attention model, a multi-head transformer encoder enabling deep bidirectional attention between modalities. Clinical and imaging embeddings were projected to a shared feature space, with cross-attention layers learning task-specific correlations.

### ResNet ANN Model Prediction

A simple fully connected artificial neural network (ANN) was constructed to incorporate race and age along with a 1-year ResNet prediction score. There are thus 3 inputs: a numerical age, a binary value (0 or 1) indicating if the participant is Black, and LDCT-based score generated by the ResNet representing the probability of cancer diagnosis. Participants listed as “more than one race” in the NLST cohort were evaluated as not Black. The ANN model has 6 outputs, each output representing the probability of lung cancer diagnosis by year N (1 to 6). There are 2 fully connected hidden layers. 100 independent ANNs were trained with Black race being weighted to equalize the proportion in the UIH cohort. Using randomly initialized parameters, the top 10 performers on a validation set (a random 10% of the UIH cohort participants) were used to create an ensemble to evaluate ResNet predictive performance on a held out test set (90% of UIH participants).

### Evaluating Model Performance

Model performance was evaluated on completely held out test sets within the NLST cohort as well as the UIH cohort. Test set IDs will be included in a Zenodo repository upon publication of the manuscript. Receiver operating characteristic (ROC) area-under-curve values generated using the Scikit-Learn Python library. To evaluate other aspects of model performance (i.e., specificity, PPV, NPV, F1) a sensitivity cutoff as close to 0.8 as possible was selected to mirror proposed sensitivities utilized in the International Lung Screening Trial for the PLCO_m2012_ model^20–22^.

### Comparison of models and performance among populations

To determine whether there was a significant difference in the ROC curve of a given model on a similar population, DeLong’s test^23^ with a modified bootstrapping approach was utilized^24^. For precision recall curve analysis on NLST White versus Black participants, 1000 times bootstrap resampling was performed. Unpaired analysis on ROC curves was performed as previously described by Hanley and McNeil^25^. P-values were adjusted for multiple hypothesis testing using Bonferroni correction.

## RESULTS

NLST and UIH cohorts evaluated in this study and racial and ethnic breakdown initially comprised of 53,452 participants in NLST and 11,654 participants in UIH (**Fig 1A-B**, **Table 1**).

**Figure 1.**
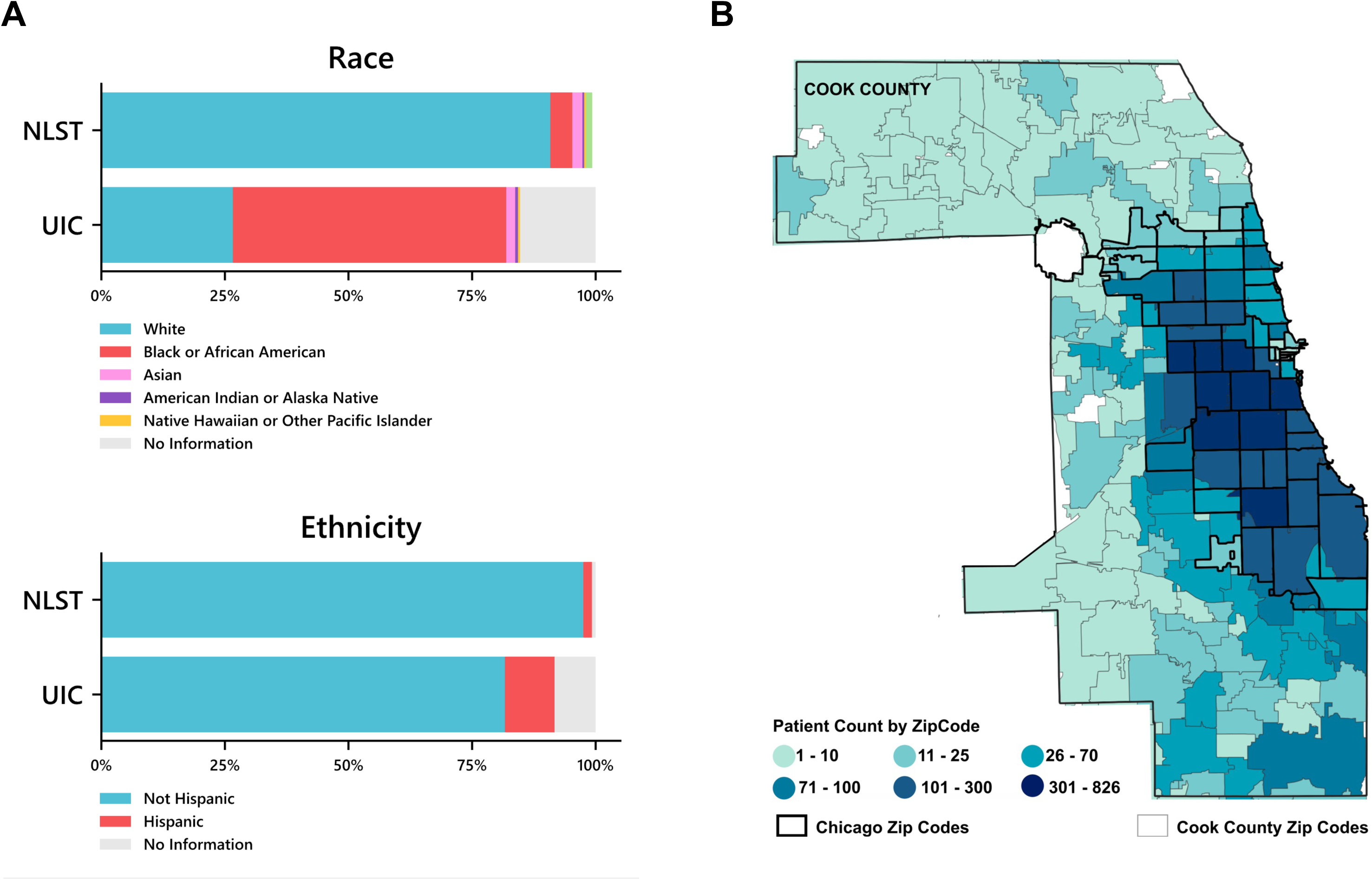
Racial and ethnic distributions in the NLST and UIH cohorts **A)** Composition of races and ethnicities in the National Lung Screening Trial (NLST) and UIH cohorts. **B)** Geographic distribution of the UIH cohort with participants counted within zip codes.

**Table 1.** Population characteristics of the NLST and UIH cohorts.

| Characteristic | NLST | UIH |
| --- | --- | --- |
| **Race** |  |  |
| White | 48,549 | 3,099 |
| Black | 2,376 | 6,447 |
| Asian | 1,095 | 210 |
| American Indian or Alaska Native | 190 | 68 |
| Native Hawaiian or Other Pacific Islander | 193 | 47 |
| Other/No Information | 1,049 | 1,783 |
| **Ethnicity** |  |  |
| Not Hispanic | 52,118 | 9,514 |
| Hispanic | 935 | 1,170 |
| No Information | 399 | 970 |
| **Lung cancer incidence** | 3.2% | 8.1% |

We sought to develop a hybrid predictive model to combine the long-term predictive potential of clinical features as well as near-term predictive potential of radiographic features from LDCT. To develop a hybrid lung cancer prediction model, we sought the most efficient means to predict lung cancer with clinical features both in terms of number and types of clinical features and model architecture. We used the clinical features from the NLST as a training data set inclusive of participants from the CT scans and Chest X-Ray arms amounting to 48,628 total participants^2^. We started first with support vector machine (SVM) and also evaluated XG Boost, a Cox Hazard implementation of XG Boost, Decision Trees, and a three layer transfomer. The SVM and other models model was chosen owing to its interpretability of features and weights and allowed for multi-year risk prognostication from baseline characteristics of participants at entry (**Figure 2A, eFigure 2**). The SVM was also selected as it is effective at classifying two classes (i.e. those with no lung cancer diagnosis versus those with lung cancer diagnosis) where ground-truth was measured by the time from study entry to cancer incidence within the window of that year’s specific model (e.g. SVM_1_ predicts the probability of lung cancer within one year or less, SVM_2_ within two years etc.). We first focused on and initiated the SVM models with 63 features available in the NLST data and utilized recursive feature elimination to optimize the model to 11 features (**eFigure 2, eTable 2**). These 11 features overlapped with many previous features identified in the PLCO_m2012_ 6-year risk model but enabled the SVM to predict multi-year risk as opposed to single 6-year risk (**eFigure 2**, **eTable 2**)^18^. We then further optimized the model for practical application by eliminating features not captured in the UIH electronic health record during routine clinical care. The excluded features included former vs current smoker, whether participants have a specific family history of lung cancer, participants’ education whether higher than bachelor’s or not, and the maximum number of years exposed to one of the following at work: asbestos, chemical, sand blasting, coal, foundry. We found that the SVM model could be reduced to a pragmatic 7 clinical feature set while still retaining predictive performance in the NLST cohort (**Fig. 2B, eTable 3**). Performance of the 7-feature multiyear SVM model was first evaluated in a held-out test set from the NLST cohort with ROC-AUC values of 0.64-0.67 similar to the 6-year lung cancer risk when compared to the PLCO_m2012_ model (**eFigure 3, eTable 4**). We then applied the model to the UIH cohort and found similar performance with ROC AUC 0.60-0.65 across years and comparable Year 6 predictive performance when compared to the PLCO_m2012_ and 11 feature SVM (**Figure 2B, eTable 4**). Except for the Transformer (ROC-AUC 0.63-0.65), other model architectures did not meet equivalence with the SVM with the exception of a three layer transformer that used all clinical features (**Data Supplement**).

**Figure 2.**
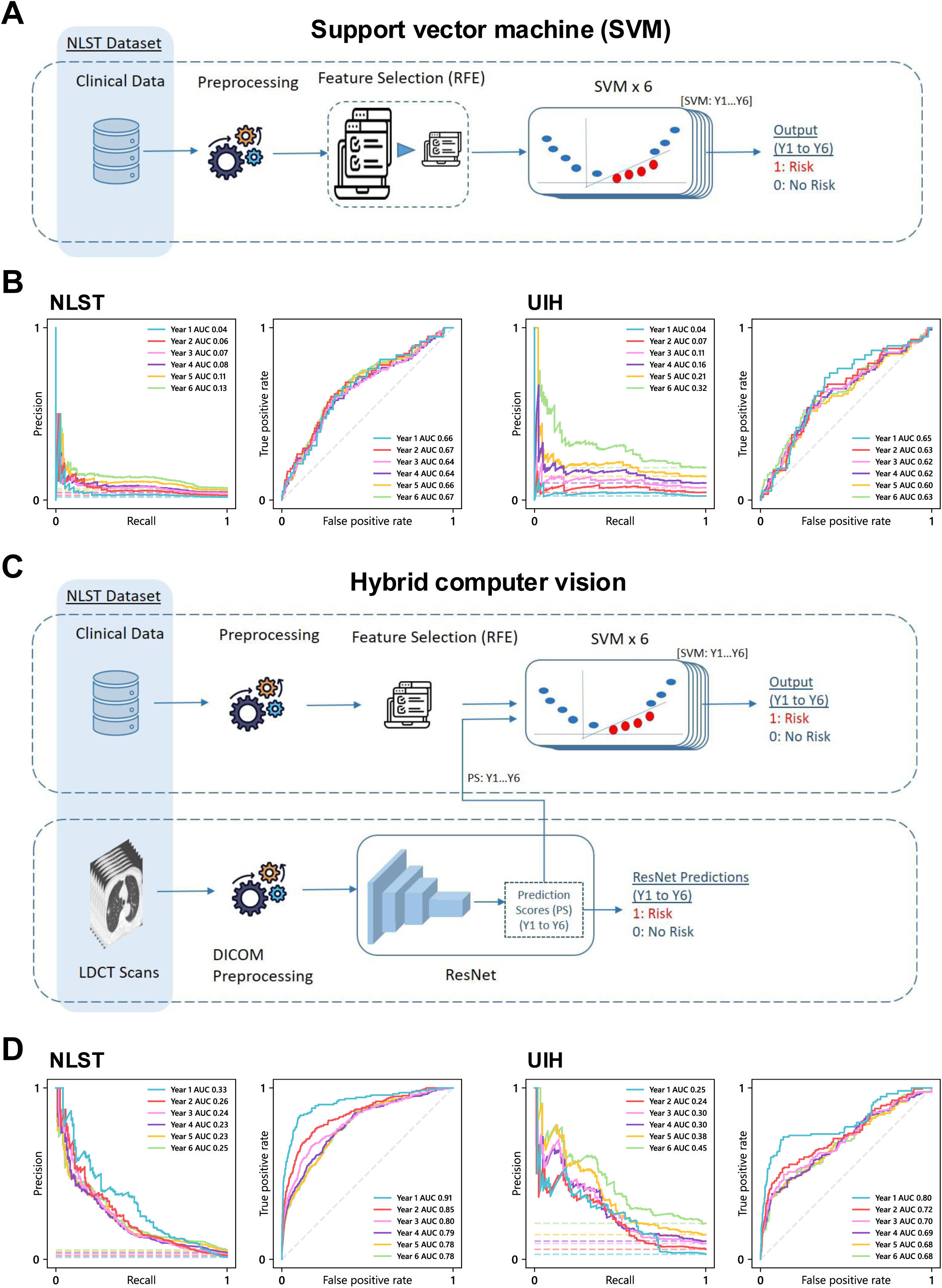
Training and testing schematics and performance of the clinical and hybrid computer vision models in the NLST and UIH cohorts. **A)** Support Vector Machine (SVM) model schema to predict lung cancer risk using NLST clinical and demographic data. **B)** 7-feature SVM performance assessed by ROC curves in the NLST (left) and UIH cohorts (right). **C)** Hybrid computer vision model (SVM + ResNet18) schema combining LDCT scans with clinical and demographic data to predict lung cancer risk. **D)** Hybrid computer vision model performance assessed by ROC curves in the NLST (left) and UIH cohorts (right).

Following initial model optimization of clinical risk features, we then incorporated an image-based lung risk predictor of the ResNet-18 CNN image classifier with additional training on LDCT images from NLST, Sybil^10^, which infers lung cancer risk probabilities at multiple years of follow up. This multimodal hybrid computer vision model was trained with the previously identified 7 clinical features in the SVM but with inclusion of ResNet model risk inference from 65,161 LDCT study DICOM files in the NLST training set (**Figure 2C**) as a late fusion strategy. We also explored additional earlier fusion strategies fusing the ResNet embeddings with a variety of cross attention architectures including Transformer, Graph Neural Networks, and a customized 2-layer cross attention model (Methods). We then benchmarked the hybrid computer vision model on the NLST test set compared to SVM models trained with clinical risk features alone which resulted in a substantial and significant improvement of predictive potential across all years (**Figure 2D**; ROC-AUC range 0.78-0.91, F1 range 0.11-0.19, **eTable 5**). Earlier fusion/cross attention architectures did not outperform the SVM hybrid computer vision model (**Data Supplement**). At a sensitivity of 80% (corresponding to a PLCO_m2012_ 6-year risk of 1.6%)^18^ we found the SVM based hybrid computer vision model had improvement in specificity when compared to the PLCO_m2012_ risk predictive model ((0.614 versus 0.411, **eTable 4**)^18^.

We then evaluated the hybrid computer vision model in the diverse UIH cohort and observed a substantial decrease in model performance (**Figure 2D, eTable 5**; ROC AUC 0.68-0.80, F1 0.017-0.238). Since we did not observe a decrease between NLST and UIH cohorts in the 7 clinical feature SVM (**Figure 2B**), we investigated the ResNet component of the Hybrid model to improve performance. To ensure ResNet model inference was reproducible in our hands, we reanalyzed individual CT scans from the NLST test set and confirmed findings as previously reported^10^ (**eFigure 4, eTable 6**). We then evaluated baseline LDCT scans using the ResNet model alone on the UIH cohort and identified significantly reduced performance in the UIH cohort across all years with Year 1 ROC-AUC values of 0.8 in UIH versus 0.92 in NLST, for example (**Table 2**, **eFigure 5**). We then asked whether model performance deterioration^26^ was due to a technical difference of the LDCT scans (e.g. modern CT scanners and image reconstruction in UIH versus NLST) or due to the demographic differences such as race. Examining the NLST data set, we did not identify any significant differences in performance when comparing scanner manufacturer or reconstruction models (**Data Supplement**). Evaluating performance of the ResNet model on White participants in the UIH showed no significant differences in performance (**Table 2**) between the NLST and the UIH cohorts. However, when assessed in Black participants, there was marked and significant deterioration in model performance with Y1-Y6 ROC AUC values 0.65-0.75 as compared to White participants in the UIH cohort (**Table 2**). Notably, ROC-AUC values in the NLST cohort were not markedly different between White and Black subjects (**Table 2**) though a low number of positive cases (e.g. 1 positive case at Year 1 in Black NLST subjects) led us to examine Precision-Recall AUC (**Table 2**) which demonstrated a significant decrease in performance in Black versus White subjects in NLST (**Table 2**). We also evaluated other potential areas of model deterioration including ethnicity, sex, and BMI and found differences in BMI where model performance had deterioration in obese individuals (BMI ≥ 30, **eTable 7**). In the NLST cohort Y1 ROC AUC was 0.94 in BMI < 30 versus 0.88 in BMI ≥ 30 subjects (**eTable 7**). In the UIH cohort, we observed more pronounced model deterioration where Y1 ROC AUC was 0.90 BMI < 30 versus 0.67 in BMI ≥ 30 subjects (**eTable 8**). These findings were also confirmed using multi-variate analysis (eSupplement).

**Table 2.**
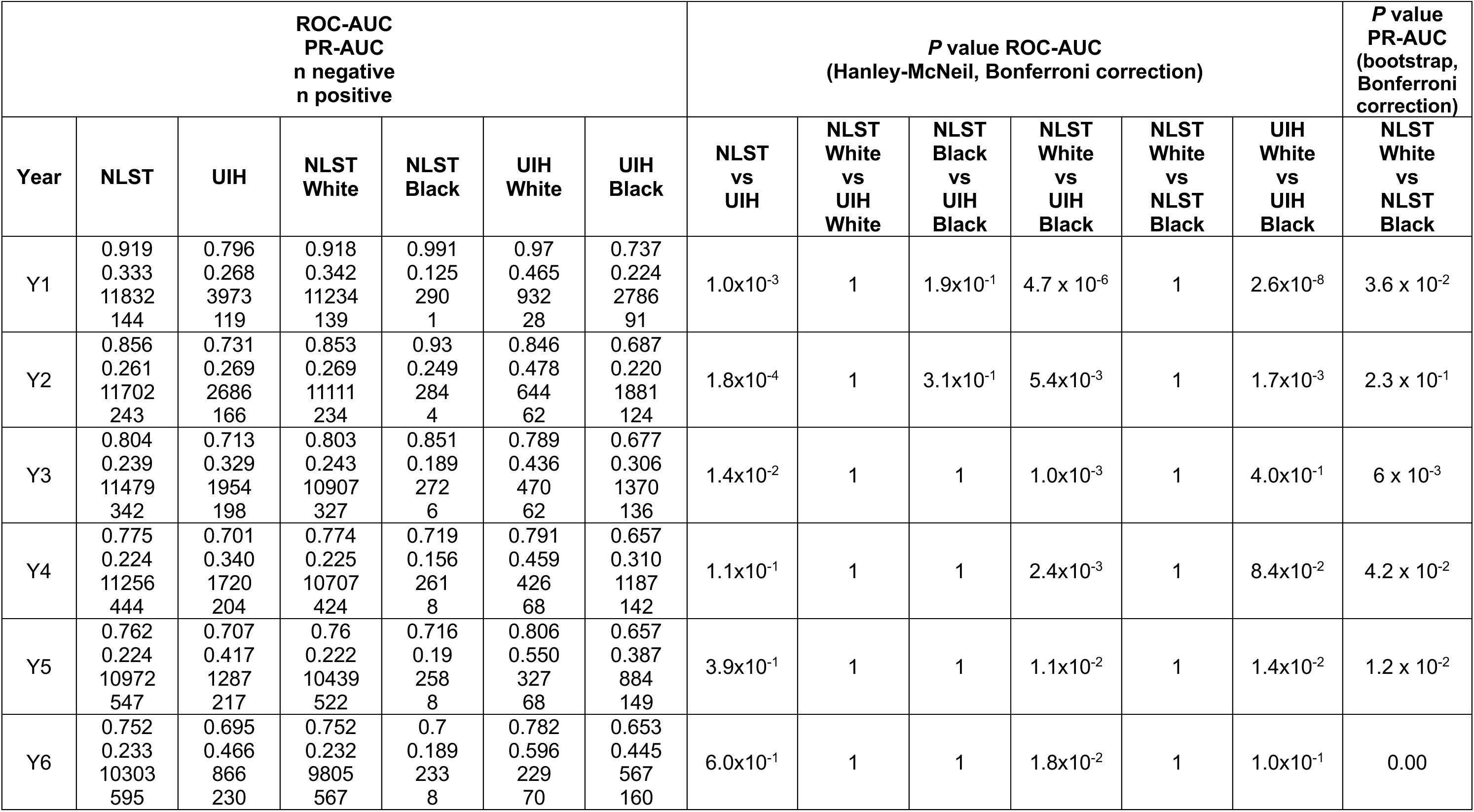
ResNet performance comparison across races in the NLST and UIH cohorts.

We then inquired whether ResNet model racial disparity could be mitigated by re-weighting the NLST cohort to increase the proportion of Black participants to that of the UIH cohort, as well as including an additional layer that incorporated race as an input. After training and testing an ensemble of Artificial Neural Networks on the NLST training set with (**eTable 9**), we evaluated the model in the UIH cohort and observed that ROC AUC values modestly increased in Black subjects by 0.01 to 0.02 across Years 1-6 as compared to the ResNet model alone (**eTable 10, Table 2**). However, in White subjects, there was a substantial decrease in performance in Y2-6 ROC-AUC values by 0.10 (**eTable 10**) as compared to the ResNet model alone (**Table 2**) potentially reflecting a domain shift or interdependencies of other clinical factors with race. In light of this domain shift and racial imbalance in the NLST cohort, we finally asked whether retraining the hybrid-computer vision model using the UIH cohort could at least improve performance in Black participants, especially given that the ResNet model did not exhibit deterioration within White UIH participants (**Table 2**). Using a 70:30 training:test split, and retraining the Hybrid Computer Vision model with the ResNet component frozen, we observed an improvement in model ROC-AUC ranging from 0.05-0.10 across years in all subjects and 0.1-0.18 in Black subjects with significance in a subset of years (**Table 3**).

**Table 3.** Predictive performance metrics of the hybrid computer vision model after retraining on 70% of the UIH cohort and testing on a 30% held out set.

|  | Year | n negative<br>n positive | ResNet<br>ROC-AUC | Retrained hybrid<br>computer vision<br>ROC-AUC | *P*<br>Bootstrapped Delong's Test<br>(Bonferroni correction) |
| --- | --- | --- | --- | --- | --- |
| All subjects | 1 | 1163<br>34 | 0.68 | 0.78 | $1.7 \times 10^{-3}$ |
| | 2 | 754<br>49 | 0.66 | 0.76 | $4.0 \times 10^{-2}$ |
| | 3 | 530<br>52 | 0.66 | 0.75 | $6.6 \times 10^{-2}$ |
| | 4 | 475<br>55 | 0.65 | 0.70 | $5.5 \times 10^{-1}$ |
| | 5 | 341<br>58 | 0.66 | 0.73 | $1.4 \times 10^{-1}$ |
| | 6 | 239<br>60 | 0.65 | 0.79 | $3.1 \times 10^{-6}$ |
| Black subjects | 1 | 751<br>28 | 0.59 | 0.67 | $9.0 \times 10^{-3}$ |
| | 2 | 475<br>43 | 0.58 | 0.68 | $8.4 \times 10^{-2}$ |
| | 3 | 337<br>46 | 0.59 | 0.74 | $1.9 \times 10^{-3}$ |
| | 4 | 289<br>49 | 0.58 | 0.70 | $2.5 \times 10^{-2}$ |
| | 5 | 173<br>52 | 0.59 | 0.70 | $1.6 \times 10^{-2}$ |
| | 6 | 123<br>54 | 0.56 | 0.74 | $2.0 \times 10^{-7}$ |
| non-Black subjects | 1 | 412<br>6 | 1 | 1 | NA |
|  | 2 | 279<br>6 | 1 | 1 | NA |
|  | 3 | 203<br>6 | 1 | 1 | NA |
|  | 4 | 186<br>6 | 1 | 1 | NA |
|  | 5 | 148<br>6 | 1 | 1 | NA |
|  | 6 | 116<br>6 | 1 | 1 | NA |

## DISCUSSION

Lung cancer disparities are extensively characterized in Black populations with respect to incidence, clinical management, screening, and survival and are associated with social determinants of health^16,17,27^. While LDCT screening has proven effective in reducing mortality through early detection and improved survival^2,18^, existing guidelines have been shown to perpetuate inequities. The USPSTF LDCT criteria^18^, primarily based on the nearly 95% White NLST study population^2^ has been shown to contribute to disparities where Black screening eligible patients carry twice the risk of lung cancer as compared to White populations^4,5^. Alternative strategies have been proposed including utilizing clinical risk models to determine screening eligibility thereby mitigating disparities of risk^5,7^. While these strategies demonstrate increased sensitivity as compared to USPSTF criteria, they have several limitations including patient selection only at long-term risk levels (> 1-1.5% lung cancer risk), lack of risk adjustment from LDCT findings, limited PPV and NPV values, and potential disparities in predictive performance (**eFigure 3**).

The hybrid computer vision model demonstrated the ability to individualize lung cancer risk in NLST and UIH cohorts, though with higher performance among populations identifying as White as compared to Black, potentially reflecting bias from an NLST training set comprised of >95% White. Bias in AI models is an increasing concern, particularly among institutions serving diverse patient populations and may further exacerbate health disparities^28,29^. However, many initiatives are aiming to reduce bias in AI development including AIM-AHEAD^30^ and specifically in lung cancer screening, the National Cancer Institute’s CANDID-4AI^23^. Because of the existing disparities in lung cancer including the higher risk of lung cancer observed among Black patients who smoke, implementation of AI models will require representative data in training sets and inclusion of Black populations and other minorities in validation cohorts and prospective studies. Furthermore, the domain shift we observed when using ANN correction of ResNet predictions may suggest that lung parenchymal features in White LDCT images possess different features and latent representations than in Black individuals, and extrapolating this with Pareto Front estimates suggest that individual versus global fairness is not optimal with the current ResNet model, especially in the context of varying percentages of Black individuals within a population (**eFigure6**). Previous reports^31,32^ emphasized the importance of population-representative training in reducing bias and improving model generalizability across racial and ethnic groups. We contextualize our race-aware and subgroup evaluations considering these findings, particularly emphasizing the goal of equitable sensitivity across demographic groups in screening scenarios. Our findings suggest that these may be overcome with retraining with more representative data (**Table 3**). In addition, we observed model deterioration in individuals with a BMI ≥ 30. While the underlying mechanism for this deterioration in both the NLST and UIH cohorts was outside of the scope of this study, we speculate that latent features within lung parenchyma may be inadequately visualized due to a biological obesity paradox that confounds our hazard risk models^33,34^. Alternatively, chest wall thickness from subcutaneous adipose tissue may interfere with ray penetration, or cause compression of lung parenchyma due to obesity hypoventilation. Further research into this phenomenon with appropriate radiology clinical domain expertise will improve generalizability through addressing the root cause(s) at image acquisition, data processing, model development, or inference^35,36^.

This study has several limitations, chiefly those of the retrospective nature of the UIH cohort which is subject to bias and data missingness. We are aiming to address these limitations with a prospective study on new LDCT participants across multiple sites. The hybrid computer vision model also utilizes a CNN architecture trained on the NLST cohort which has limited representation of Black and other minoritized populations. Current efforts such as the CANDID-4AI initiative^37^ to develop more inclusive training sets would enable new models to overcome the domain shifts we observed in the ResNet model^10^ where performance in White subjects decreased when the model was optimized for Black subjects. Having inclusive and representative training sets with race labeling will allow ResNet and newer foundation models to have greater generalizability and understand underlying features of lung cancer racial disparities. In addition to more diverse training sets, domain adaptation and adversarial learning to debias the NLST cohort are approaches we did not attempt but could also be investigated^35,36,38^. Finally, when retraining the hybrid computer vision model on the UIH population, we intentionally left the ResNet layer frozen to avoid adapting the vision component to internal variance of imaging hardware/software at UIH. Nevertheless, our results may still represent overfitting to either physical or geographic factors in UIH participants as well as site-specific artifacts or sociotechnical confounders. To truly evaluate model performance and generalizability, a similarly diverse but a geographically distinct cohort of LDCT subjects would be required for both training and testing. Another limitation is that our study focuses on conventional static, snapshot-based prediction of future lung cancer risk at fixed time horizons, as opposed to more modern architectures designed for time to event series detection^39,40^. An important direction for future research would incorporate these approaches to model the temporal progression of clinically significant lung cancer, especially with the incorporation of longitudinal data. As more temporally aligned LDCT scans and clinical records become available, there is a valuable opportunity to incorporate time series modeling techniques. We consider this an exciting and necessary extension of the current work as longitudinal datasets mature.

A recent development in the field of medical artificial intelligence are vision language models including frontier models such as MedGemma and MedSigLip^41^ which are trained foundation models on medical images and text, and have strong potential to outperform ResNet based architectures trained on more generalized imaging data sets. While this study does not incorporate vision-language models (vLLMs) directly, we recognize that such models, as well as multimodal learning systems more broadly, have been shown to reflect and potentially amplify biases present in their training data. Recent work has highlighted demographic, linguistic, and cultural biases in LLMs^29^, which can adversely affect clinical decision support systems if left unchecked. As the field increasingly moves toward multimodal diagnostic frameworks, including those integrating textual clinical notes and medical images, it will be critical to assess and mitigate bias at the intersection of modalities. Future extensions of this work may explore the integration of language-based data, with attention to bias auditing and subgroup performance.

Overall, we demonstrate that an integrated approach combining imaging and clinical features can predict lung cancer risk at an individual level, and that model deterioration in Black populations can potentially be overcome by retraining on more inclusive and representative cohorts. Further validation of this approach and similar AI tools can assist clinicians in managing risk reduction and preventive measures by estimating future lung cancer risk and identifying high risk individuals. Lastly, accurate and equitable lung cancer risk assessments can enable personalized initiation of LDCT screening which in turn may mitigate racial disparities in lung cancer screening and survival.

## Data Availability

All data and code used to generate figures and tables in this manuscript will be made available in a Zenodo repository upon the publication of this manuscript. Sharing of individual level deidentified data is subject to review and approval of the University of Illinois Chicago Institutional Review Board and will be distributed following the University of Illinois Chicago INDIGO repository standard practices under a Material Transfer Agreement.

## ACKNOWLEDGEMENTS

We wish to thank the UIC Center for Clinical Translational Sciences for technical support, the UI Cancer Center Data Integrated Shared Resource (DISR) for their provision of population-level data and data visualization/mapping services., J.R., M.M.P, and F.D.W. for helpful discussions on the manuscript. A.J.Z. was supported by an AI.Health4all postdoctoral fellowship, M.B. was supported by a Winn-CIPP fellowship, A.A.S. received support as a National Cancer Institute, Center for Cancer Health Equity Early Investigator Advancement Program Scholar. A.J.Z., N.P., and A.A.S. conducted the data analysis presented in this manuscript. A.A.S. had full access to all the data in the study and takes responsibility for the integrity of the data and the accuracy of the data analysis.

## CONFLICTS OF INTEREST

A.A.S. and A.J.Z. have filed an invention disclosure related to this work. A.A.S. was recently employed by Tempus AI., Inc. and holds an equity interest. A.A.K. and V.K.G. have equity interests in Tempus AI.

## Notes

### Funding Statement

This study was funded by the UIC AI.Health4All Center for Health Equity using Machine Learning and Artificial Intelligence.

### Author Declarations

IRB of University of Illinois at Chicago gave ethical approval for this work under protocol numbers listed in this manuscript.

### Summary of Updates

Additional results and analyses have been added.

## REFERENCES

1. Soneji S, Tanner NT, Silvestri GA, et al: Racial and Ethnic Disparities in Early-Stage Lung Cancer Survival. Chest 152:587–597, 2017

2. National Lung Screening Trial Research T, Aberle DR, Adams AM, et al: Reduced lung-cancer mortality with low-dose computed tomographic screening. N Engl J Med 365:395–409, 2011

3. Bandi P, Star J, Ashad-Bishop K, et al: Lung Cancer Screening in the US, 2022. JAMA Intern Med, 2024

4. Pinsky PF, Lau YK, Doubeni CA: Potential Disparities by Sex and Race or Ethnicity in Lung Cancer Screening Eligibility Rates. Chest 160:341–350, 2021

5. Aredo JV, Choi E, Ding VY, et al: Racial and Ethnic Disparities in Lung Cancer Screening by the 2021 USPSTF Guidelines Versus Risk-Based Criteria: The Multiethnic Cohort Study. JNCI Cancer Spectr 6, 2022

6. Choi E, Ding VY, Luo SJ, et al: Risk Model-Based Lung Cancer Screening and Racial and Ethnic Disparities in the US. JAMA Oncol, 2023

7. Pasquinelli MM, Tammemagi MC, Kovitz KL, et al: Risk Prediction Model Versus United States Preventive Services Task Force Lung Cancer Screening Eligibility Criteria: Reducing Race Disparities. J Thorac Oncol 15:1738–1747, 2020

8. Field JK, Vulkan D, Davies MPA, et al: Liverpool Lung Project lung cancer risk stratification model: calibration and prospective validation. Thorax 76:161–168, 2021

9. Katki HA, Kovalchik SA, Berg CD, et al: Development and Validation of Risk Models to Select Ever-Smokers for CT Lung Cancer Screening. JAMA 315:2300–11, 2016

10. Mikhael PG, Wohlwend J, Yala A, et al: Sybil: A Validated Deep Learning Model to Predict Future Lung Cancer Risk From a Single Low-Dose Chest Computed Tomography. J Clin Oncol 41:2191–2200, 2023

11. Heuvelmans MA, van Ooijen PMA, Ather S, et al: Lung cancer prediction by Deep Learning to identify benign lung nodules. Lung Cancer 154:1–4, 2021

12. Ardila D, Kiraly AP, Bharadwaj S, et al: End-to-end lung cancer screening with three-dimensional deep learning on low-dose chest computed tomography. Nat Med 25:954–961, 2019

13. Liao F, Liang M, Li Z, et al: Evaluate the Malignancy of Pulmonary Nodules Using the 3-D Deep Leaky Noisy-OR Network. IEEE Trans Neural Netw Learn Syst 30:3484–3495, 2019

14. Centers for Disease C, Prevention: Racial/Ethnic disparities and geographic differences in lung cancer incidence --- 38 States and the District of Columbia, 1998-2006. MMWR Morb Mortal Wkly Rep 59:1434–8, 2010

15. Byrne CA, Gomez SL, Kim S, et al: Disparities in inflammation between non-Hispanic black and white individuals with lung cancer in the Greater Chicago Metropolitan area. Front Immunol 13:1008674, 2022

16. Watson KS, Hulbert A, Henderson V, et al: Lung Cancer Screening and Epigenetics in African Americans: The Role of the Socioecological Framework. Front Oncol 9:87, 2019

17. Kim SJ, Kery C, An J, et al: Racial/Ethnic disparities in exposure to neighborhood violence and lung cancer risk in Chicago. Soc Sci Med 340:116448, 2024

18. Tammemagi MC, Katki HA, Hocking WG, et al: Selection criteria for lung-cancer screening. N Engl J Med 368:728–36, 2013

19. Pedregosa F, Varoquaux G, Gramfort A, et al: Scikit-learn: Machine learning in Python. the Journal of machine Learning research 12:2825–2830, 2011

20. Weber M, Yap S, Goldsbury D, et al: Identifying high risk individuals for targeted lung cancer screening: Independent validation of the PLCO(m2012) risk prediction tool. Int J Cancer 141:242–253, 2017

21. Tammemagi MC, Ruparel M, Tremblay A, et al: USPSTF2013 versus PLCOm2012 lung cancer screening eligibility criteria (International Lung Screening Trial): interim analysis of a prospective cohort study. Lancet Oncol 23:138–148, 2022

22. Force USPST, Krist AH, Davidson KW, et al: Screening for Lung Cancer: US Preventive Services Task Force Recommendation Statement. JAMA 325:962–970, 2021

23. DeLong ER, DeLong DM, Clarke-Pearson DL: Comparing the areas under two or more correlated receiver operating characteristic curves: a nonparametric approach. Biometrics 44:837–45, 1988

24. Demler OV, Pencina MJ, D’Agostino RB, Sr.: Misuse of DeLong test to compare AUCs for nested models. Stat Med 31:2577–87, 2012

25. Hanley JA, McNeil BJ: The meaning and use of the area under a receiver operating characteristic (ROC) curve. Radiology 143:29–36, 1982

26. Zhang A, Xing L, Zou J, et al: Shifting machine learning for healthcare from development to deployment and from models to data. Nat Biomed Eng 6:1330–1345, 2022

27. Kim SJ, Peterson CE, Warnecke R, et al: The Uneven Distribution of Medically Underserved Areas in Chicago. Health Equity 4:556–564, 2020

28. Crowe B, Rodriguez JA: Identifying and Addressing Bias in Artificial Intelligence. JAMA Netw Open 7:e2425955, 2024

29. Liu Z: Cultural bias in large language models: A comprehensive analysis and mitigation strategies. Journal of Transcultural Communication 3:224–244, 2025

30. Hendricks-Sturrup R, Simmons M, Anders S, et al: Developing Ethics and Equity Principles, Terms, and Engagement Tools to Advance Health Equity and Researcher Diversity in AI and Machine Learning: Modified Delphi Approach. JMIR AI 2:e52888, 2023

31. Etzel CJ, Kachroo S, Liu M, et al: Development and validation of a lung cancer risk prediction model for African-Americans. Cancer Prev Res (Phila) 1:255–65, 2008

32. Landy R, Gomez I, Caverly TJ, et al: Methods for Using Race and Ethnicity in Prediction Models for Lung Cancer Screening Eligibility. JAMA Netw Open 6:e2331155, 2023

33. Cespedes Feliciano EM, Kroenke CH, Caan BJ: The Plausibility of the Obesity Paradox in Cancer-Response-Reply to Point. Cancer Res 78:1904–1905, 2018

34. Park Y, Peterson LL, Colditz GA: The Plausibility of Obesity Paradox in Cancer-Point. Cancer Res 78:1898–1903, 2018

35. Kim W: Seeing the Unseen: Advancing Generative AI Research in Radiology. Radiology 311:e240935, 2024

36. Banerjee I, Bhattacharjee K, Burns JL, et al: "Shortcuts" Causing Bias in Radiology Artificial Intelligence: Causes, Evaluation, and Mitigation. J Am Coll Radiol 20:842–851, 2023

37. Institute NC: Leveraging Artificial Intelligence to Improve Accuracy of Lung Cancer Screening was originally published by the National Cancer Institute., 2022

38. Yang J, Soltan AAS, Eyre DW, et al: An adversarial training framework for mitigating algorithmic biases in clinical machine learning. NPJ Digit Med 6:55, 2023

39. Li J, Fearnhead P, Fryzlewicz P, et al: Automatic change-point detection in time series via deep learning. submitted. arXiv preprint arxiv:2211.03860, 2022

40. Azib M, Renard B, Garnier P, et al: Event detection in time series: universal deep learning approach. arXiv preprint arXiv:2311.15654, 2023

41. Golden DaP, R: MedGemma: Our most capable open models for health AI development. Google Research Blog, Google Research, 2025

