## Supplemental Material for "A hybrid computer vision model to predict lung cancer in diverse populations"

**eTable 1 – Inclusion/exclusion criteria for participants in the retrospective cohort of UIH**

| Criteria | Description |
|---|---|
| Time | January 1, 2015 – March 16, 2024 |
| Age | 50-80 as of March 16, 2024 |
| Tobacco | ICD-9 305.1; ICD-10, F17; history of smoking, HCPCS:G0297,CPT4:71271, |
| CT chest | CPT 87.4100, 87.41, 87.4102, 87.4200, 87.4101, 87.42, BP2YZZZ, BP2YYZZ, BP2Y1ZZ, BP2Y0ZZ, BP2XZZZ, BP2XYZZ, BP2X1ZZ, BP2X0ZZ, BP2WYZZ, BP2W1ZZ, BP2W0ZZ, 64490, 8E0WXBG, BW25ZZZ, BW25YZZ, BW25Y0Z, BW251ZZ, BW2510Z, BW250ZZ, BW2500Z, BW24ZZZ, BW24YZZ, BW24Y0Z, BW241ZZ, BW2410Z, BW240ZZ, BW2400Z, BR27ZZZ, BR27YZZ, BR271ZZ, BR270ZZ, B027ZZZ, B027YZZ, B027Y0Z, B0271ZZ, B02710Z, B0270ZZ, B02700Z, BB291ZZ, BB2910Z, BB290ZZ, BB2900Z, BB28ZZZ, BB28YZZ, BB28Y0Z, BB281ZZ, BB2810Z, BB280ZZ, BB2800Z, BB27ZZZ, BB27YZZ, BB27Y0Z, BB271ZZ, BB2710Z, BB270ZZ, BB2700Z, BB24ZZZ, BB24YZZ, BB24Y0Z, BB241ZZ, BB2410Z, BB240ZZ, BB2400Z, BB29ZZZ, BB29YZZ, BB29Y0Z, B320YZZ, B3201ZZ, B3200ZZ, B52SZZZ, B52SZ2Z, B52SYZZ, B52SY0Z, B52S1ZZ, B52S10Z, B52S0ZZ, B52S00Z, B52RZZZ, B52RZ2Z, B52RYZZ, B52RY0Z, B52R1ZZ, B52R10Z, B52R0ZZ, B52R00Z, B52QZZZ, B52QZ2Z, B52QYZZ, B52QY0Z, B52Q1ZZ, B52Q10Z, B52Q0ZZ, B52Q00Z, B529ZZZ, B529Z2Z, B529YZZ, B529Y0Z, B5291ZZ, B52910Z, B5290ZZ, B52900Z, B528ZZZ, B528Z2Z, B528YZZ, B528Y0Z, B5281ZZ, B52810Z, B5280ZZ, B52800Z, 78816, 78815, 78814, 72130, 72129, 72128, 71275, 71270, 71260, 71250 |
| No cancer history | Prior to first imaging study, no ICD9/10 code of neoplasm 149-23/C exception for non-melanoma skin cancers (ICD 9 173.X; ICD 10 C44.X) |

**eFigure 1 – Flowchart schema of LDCT DICOM filtering in UIH cohort**

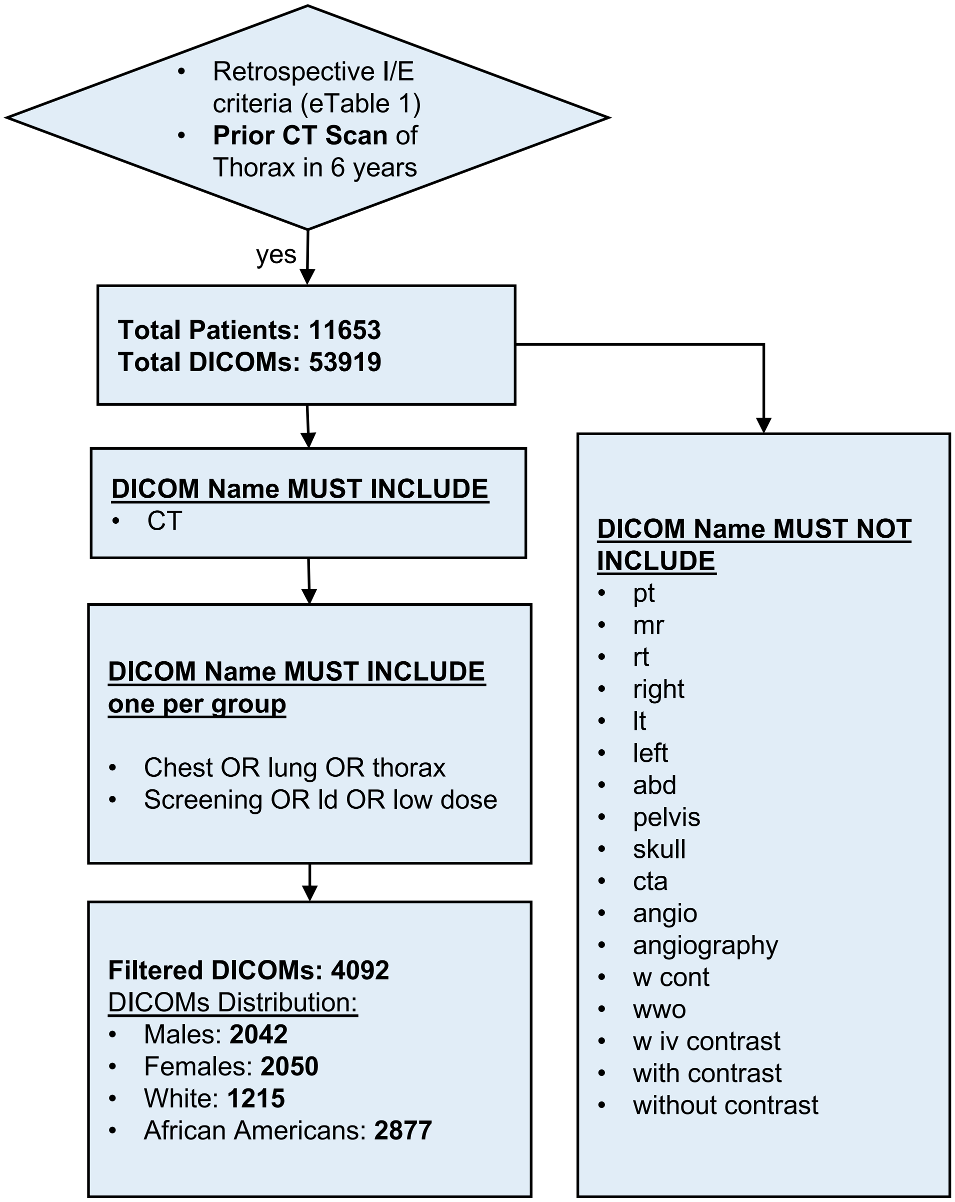

**eFigure 1.** Flowchart schema of LDCT DICOM filtering in UIH cohort. After inclusion/exclusion criteria, 11,653 participants were filtered for LDCT studies using the above schema.

### eFigure 2 – Support Vector Machine (SVM) training and testing schema

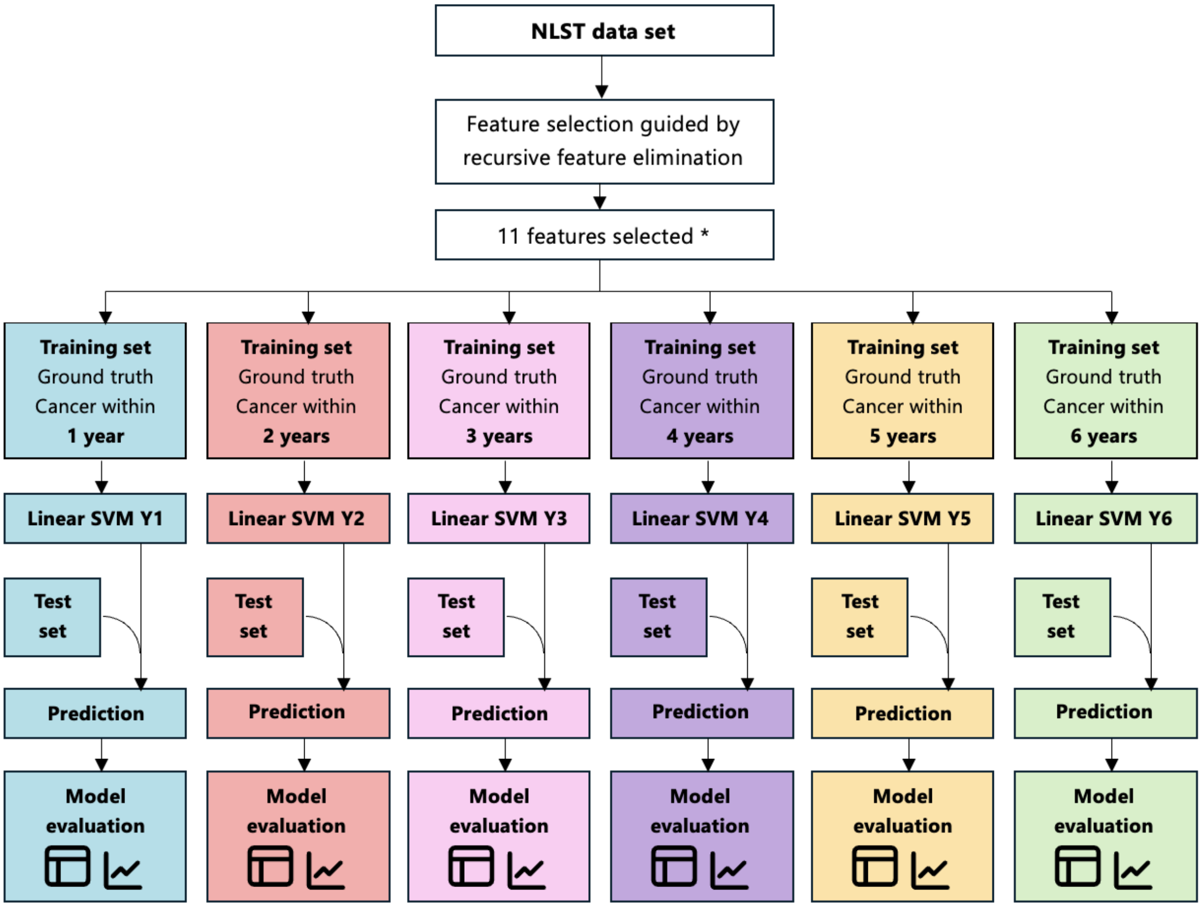

**eFigure 2.** Support Vector Machine (SVM) training and testing schema. NLST participant data inclusive of CT and chest X ray arms utilized spanned demographics, vitals, and questionnaire responses. Recursive feature elimination narrowed the feature set to eleven features (see **eTable 2**) and were used to train six independent SVMs to predict lung cancer risk for each year after NLST study entry.

**eTable 2 – Features and coefficients of 11 and 7 feature SVM models trained on NLST cohort**

| SVM11 Features | Coefficient |
|---|---|
| Smoking years | 0.551 |
| Age | 0.309 |
| Cigarettes per day | 0.283 |
| BMI | -0.229 |
| Divorced or widowed | 0.140 |
| Smoking status | 0.132 |
| Education: Bachelor's and above | -0.127 |
| Race: Black or African American | 0.120 |
| Family history of lung cancer | 0.102 |
| Ethnicity | -0.09 |
| Work exposure - asbestos, chemical, sand blasting, coal mine, or foundry | 0.06 |

| SVM7 Features | Coefficient |
|---|---|
| Smoking years | 0.67235175 |
| Cigarettes per day | 0.31821073 |
| Age | 0.24901165 |
| BMI | -0.2328077 |
| Divorced or widowed | 0.13950669 |
| Race: Black or African American | 0.10485424 |
| Ethnicity | -0.0914769 |

### eTable 3 - Bootstrap resampling based DeLong's Statistics of 7 feature SVM vs 11 feature SVM

| Year | P-Value with Bonferroni Correction | DeLong's test statistic |
|---|---|---|
| Y1 | 1.00 | -0.07 |
| Y2 | 1.00 | 0.24 |
| Y3 | 0.30 | 1.956 |
| Y4 | 0.024 | 2.891 |
| Y5 | 0.516 | 1.718 |
| Y6 | 1.00 | -0.456 |

**eFigure 3 – PR and ROC plots of $PLCO_{m2012}$ risk prediction in the NLST test set at Year 6**

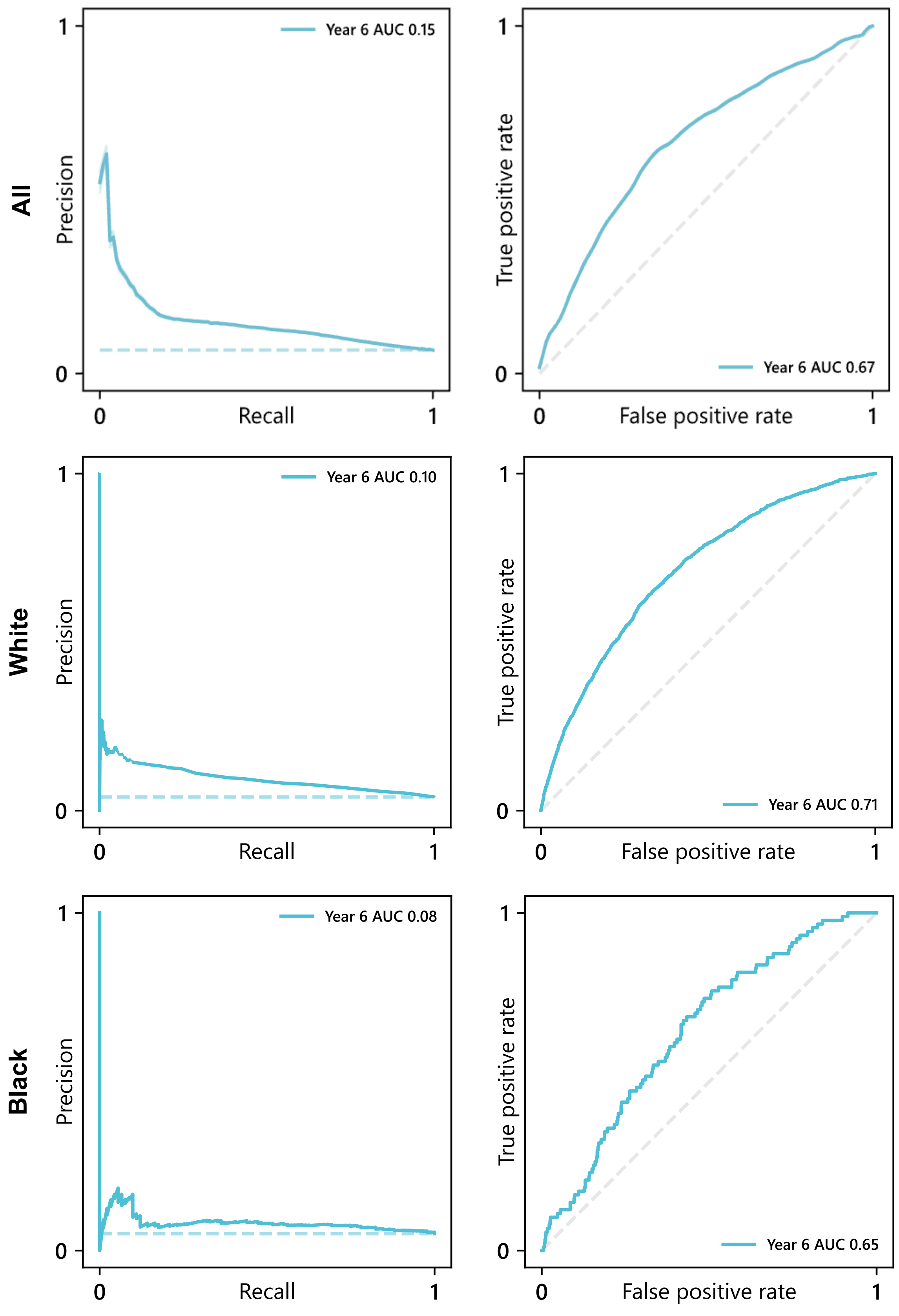

**eFigure 3.** $PLCO_{m2012}$ model prediction of lung cancer at year 6 within the NLST test set among all (top), White (middle) and Black (bottom) participants. Precision Recall curves (left) with AUC values of 0.15, 0.10, and 0.08, respectively. ROC curves (right) with AUC values of 0.67, 0.71, and 0.65, respectively.

**eTable 4 – Benchmark metrics of $PLCO_{m2012}$, SVM, hybrid computer vision (Hybrid CV) on NLST participants at Year 6**

| Year 6 Model Results | ROC AUC | PR AUC | Sensitivity | Specificity | PPV | NPV | n positive | n negative |
|---|---|---|---|---|---|---|---|---|
| $PLCO_{m2012}$ Y6 | 0.671 | 0.146 | 0.797 | 0.411 | 0.09 | 0.965 | 138 | 1896 |
| 11 feat. SVM Y6 | 0.671 | 0.137 | 0.797 | 0.4 | 0.088 | 0.964 | 138 | 1896 |
| 7 feat. SVM Y6 | 0.671 | 0.135 | 0.797 | 0.445 | 0.095 | 0.968 | 138 | 1896 |
| Hybrid CV Y6 | 0.782 | 0.246 | 0.8 | 0.614 | 0.107 | 0.982 | 595 | 10303 |

**eTable 5 – ROC-AUC metrics of Hybrid computer vision versus 7 feature SVM in the NLST and UIH cohorts and ROC-AUC statistical testing (DeLong's test with bootstrapping)**

| Cohort | Year | 7 feature SVM ROC-AUC | Hybrid computer vision ROC-AUC | *P* value with Bonferroni Correction | DeLong's test statistic | Hybrid computer vision F1 |
|---|---|---|---|---|---|---|
| NLST | 1 | 0.66 | 0.91 | 0.000 | 11.524 | .173 |
| | 2 | 0.67 | 0.85 | 0.000 | 10.231 | .109 |
| | 3 | 0.64 | 0.80 | 0.000 | 9.921 | .110 |
| | 4 | 0.64 | 0.79 | 0.000 | 9.56 | .140 |
| | 5 | 0.66 | 0.78 | 0.000 | 9.548 | .170 |
| | 6 | 0.67 | 0.78 | 0.000 | 8.929 | .186 |
| UIH | 1 | 0.653 | 0.797 | 0.000 | 9.36 | .078 |
| | 2 | 0.629 | 0.724 | 0.000 | 9.013 | .136 |
| | 3 | 0.619 | 0.696 | 0.000 | 8.918 | .207 |
| | 4 | 0.617 | 0.686 | 3.33067E-14 | 8.03 | .238 |
| | 5 | 0.598 | 0.681 | 9.32586E-15 | 8.158 | .017 |
| | 6 | 0.629 | 0.684 | 1.98508E-13 | 7.395 | .021 |

### eFigure 4 – Reproduction of ResNet model ROC curve and AUC in NLST cohort

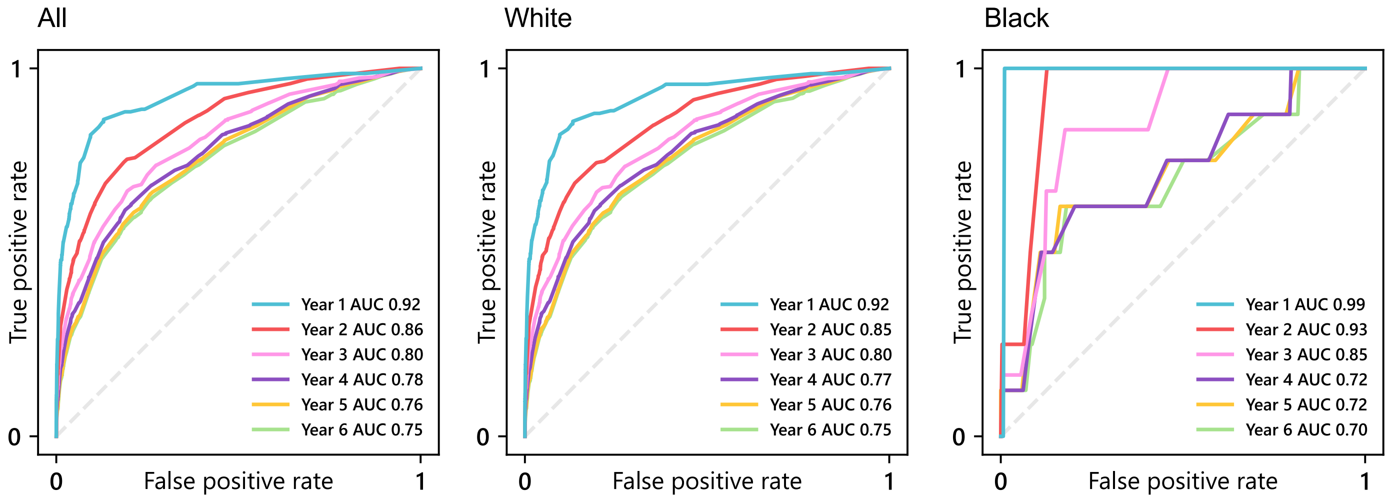

**eFigure 4 –** Reproduction of ResNet model ROC curve and AUC on NLST test set in the entire cohort (left), White participants (middle), and Black participants (right).

### eTable 6 – ResNet performance metrics of all races within the NLST cohort

| | Year | ROC AUC | PR AUC | Sensitivity | Specificity | PPV | NPV | F1 | n positive | n negative |
|---|---|---|---|---|---|---|---|---|---|---|
| All Races | Year 1 | 0.919 | 0.333 | 0.812 | 0.907 | 0.096 | 0.997 | 0.172 | 144 | 11832 |
| | Year 2 | 0.856 | 0.261 | 0.757 | 0.784 | 0.068 | 0.994 | 0.125 | 243 | 11702 |
| | Year 3 | 0.804 | 0.239 | 0.807 | 0.606 | 0.057 | 0.991 | 0.107 | 342 | 11479 |
| | Year 4 | 0.775 | 0.224 | 0.82 | 0.544 | 0.066 | 0.987 | 0.123 | 444 | 11256 |
| | Year 5 | 0.762 | 0.224 | 0.803 | 0.539 | 0.08 | 0.982 | 0.145 | 547 | 10972 |
| | Year 6 | 0.752 | 0.233 | 0.792 | 0.538 | 0.09 | 0.978 | 0.162 | 595 | 10303 |
| White | Year 1 | 0.918 | 0.342 | 0.813 | 0.905 | 0.096 | 0.997 | 0.172 | 139 | 11234 |
| | Year 2 | 0.853 | 0.269 | 0.846 | 0.647 | 0.048 | 0.995 | 0.091 | 234 | 11111 |
| | Year 3 | 0.803 | 0.243 | 0.807 | 0.605 | 0.058 | 0.991 | 0.108 | 327 | 10907 |
| | Year 4 | 0.774 | 0.225 | 0.823 | 0.543 | 0.067 | 0.987 | 0.123 | 424 | 10707 |
| | Year 5 | 0.76 | 0.222 | 0.805 | 0.538 | 0.08 | 0.982 | 0.146 | 522 | 10439 |
| | Year 6 | 0.752 | 0.232 | 0.794 | 0.538 | 0.09 | 0.978 | 0.162 | 567 | 9805 |
| Black | Year 1 | 0.991 | 0.125 | 1 | 0.99 | 0.25 | 1 | 0.073 | 1 | 290 |
| | Year 2 | 0.93 | 0.249 | 1 | 0.873 | 0.1 | 1 | 0.141 | 4 | 284 |
| | Year 3 | 0.851 | 0.189 | 0.833 | 0.824 | 0.094 | 0.996 | 0.183 | 6 | 272 |
| | Year 4 | 0.719 | 0.156 | 0.75 | 0.544 | 0.048 | 0.986 | 0.208 | 8 | 261 |
| | Year 5 | 0.716 | 0.19 | 0.75 | 0.539 | 0.048 | 0.986 | 0.270 | 8 | 258 |
| | Year 6 | 0.7 | 0.189 | 0.75 | 0.498 | 0.049 | 0.983 | 0.389 | 8 | 233 |

**eFigure 5 – ResNet model ROC curve and AUC on UIH cohort**

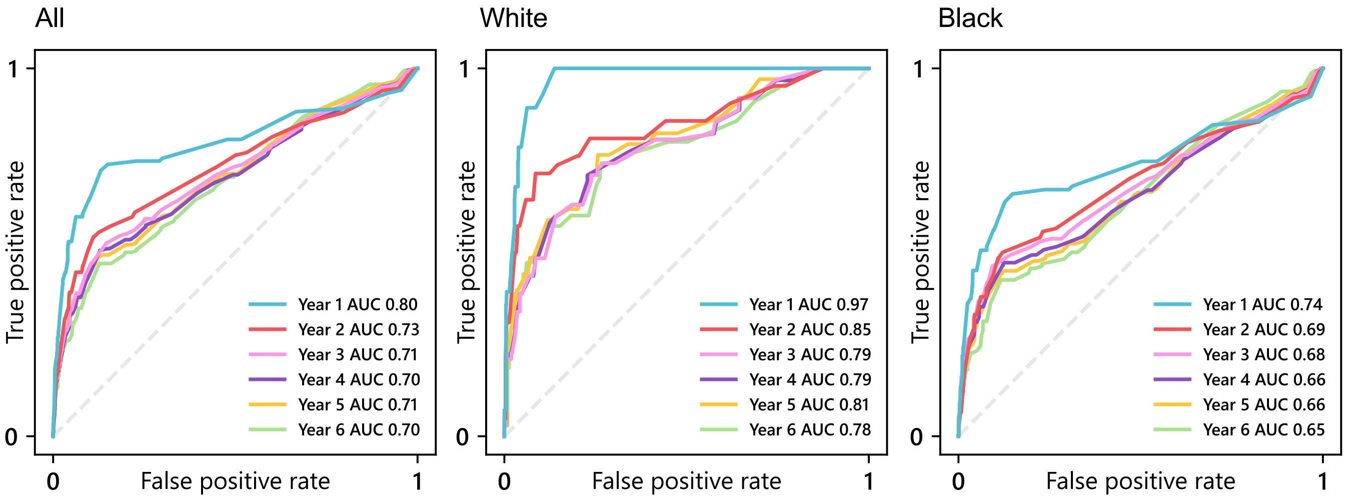

**eFigure 5 –**ResNet model ROC curve and AUC on UIH cohort among all races (left), White participants (middle), and Black participants (right).

**eTable 7 – ResNet performance metrics among individuals with varying BMI in the NLST cohort**

| Year | ROC AUC | | PR AUC | | Sensitivity | | Specificity | | PPV | | NPV | | n positive | | n negative | |
|---|---|---|---|---|---|---|---|---|---|---|---|---|---|---|---|---|
| | BMI < 30 | BMI ≥ 30 | BMI < 30 | BMI ≥ 30 | BMI < 30 | BMI ≥ 30 | BMI < 30 | BMI ≥ 30 | BMI < 30 | BMI ≥ 30 | BMI < 30 | BMI ≥ 30 | BMI < 30 | BMI ≥ 30 | BMI < 30 | BMI ≥ 30 |
| 1 | 0.94 | 0.88 | 0.37 | 0.23 | 0.78 | 0.79 | 0.92 | 0.83 | 0.11 | 0.05 | 0.99 | 0.99 | 102 | 42 | 8295 | 3537 |
| 2 | 0.87 | 0.82 | 0.29 | 0.18 | 0.80 | 0.81 | 0.80 | 0.63 | 0.08 | 0.04 | 0.99 | 0.99 | 176 | 67 | 8202 | 3500 |
| 3 | 0.82 | 0.76 | 0.26 | 0.17 | 0.82 | 0.80 | 0.63 | 0.56 | 0.06 | 0.05 | 0.99 | 0.99 | 243 | 99 | 8033 | 3446 |
| 4 | 0.78 | 0.77 | 0.23 | 0.19 | 0.78 | 0.81 | 0.59 | 0.56 | 0.07 | 0.06 | 0.99 | 0.99 | 326 | 118 | 7842 | 3414 |
| 5 | 0.76 | 0.78 | 0.23 | 0.22 | 0.80 | 0.82 | 0.53 | 0.56 | 0.09 | 0.07 | 0.98 | 0.99 | 415 | 132 | 7654 | 3318 |
| 6 | 0.74 | 0.79 | 0.23 | 0.25 | 0.82 | 0.77 | 0.44 | 0.64 | 0.09 | 0.09 | 0.98 | 0.98 | 455 | 140 | 7200 | 3103 |

**eTable 8 – ResNet performance metrics among individuals with varying BMI in the UIH cohort**

| Year | ROC AUC | | PR AUC | | Sensitivity | | Specificity | | PPV | | NPV | | n positive | | n negative | |
|---|---|---|---|---|---|---|---|---|---|---|---|---|---|---|---|---|
| | BMI < 30 | BMI ≥ 30 | BMI < 30 | BMI ≥ 30 | BMI < 30 | BMI ≥ 30 | BMI < 30 | BMI ≥ 30 | BMI < 30 | BMI ≥ 30 | BMI < 30 | BMI ≥ 30 | BMI < 30 | BMI ≥ 30 | BMI < 30 | BMI ≥ 30 |
| 1 | 0.90 | 0.67 | 0.31 | 0.22 | 0.77 | 0.78 | 0.89 | 0.26 | 0.16 | 0.03 | 0.99 | 0.97 | 65 | 54 | 2261 | 1712 |
| 2 | 0.83 | 0.59 | 0.30 | 0.23 | 0.85 | 0.76 | 0.61 | 0.12 | 0.12 | 0.05 | 0.99 | 0.89 | 94 | 72 | 1492 | 1194 |
| 3 | 0.81 | 0.57 | 0.38 | 0.30 | 0.81 | 0.77 | 0.62 | 0.16 | 0.20 | 0.07 | 0.97 | 0.89 | 120 | 78 | 1046 | 908 |
| 4 | 0.79 | 0.56 | 0.39 | 0.31 | 0.79 | 0.80 | 0.61 | 0.12 | 0.21 | 0.09 | 0.96 | 0.86 | 123 | 81 | 931 | 789 |
| 5 | 0.81 | 0.56 | 0.52 | 0.33 | 0.80 | 0.81 | 0.64 | 0.13 | 0.30 | 0.12 | 0.94 | 0.83 | 132 | 85 | 685 | 602 |
| 6 | 0.78 | 0.56 | 0.56 | 0.37 | 0.81 | 0.81 | 0.59 | 0.15 | 0.38 | 0.17 | 0.91 | 0.79 | 145 | 85 | 465 | 401 |

**eTable 9 – Artificial Neural Network retrained ResNet model metrics on all races within the NLST cohort**

| Year | ROC AUC | PR AUC | Sensitivity | Specificity | PPV | NPV | n positive | n negative |
|---|---|---|---|---|---|---|---|---|
| Year 1 | 0.904 | 0.326 | 0.799 | 0.91 | 0.098 | 0.997 | 144 | 11832 |
| Year 2 | 0.846 | 0.258 | 0.794 | 0.713 | 0.054 | 0.994 | 243 | 11702 |
| Year 3 | 0.804 | 0.238 | 0.795 | 0.658 | 0.065 | 0.991 | 342 | 11479 |
| Year 4 | 0.783 | 0.226 | 0.804 | 0.626 | 0.078 | 0.988 | 444 | 11256 |
| Year 5 | 0.771 | 0.229 | 0.803 | 0.607 | 0.092 | 0.984 | 547 | 10972 |
| Year 6 | 0.771 | 0.24 | 0.8 | 0.64 | 0.114 | 0.982 | 595 | 10303 |

**eTable 10 – Artificial Neural Network retrained ResNet model metrics on the UIH cohort**

| | Year | ROC AUC | PR AUC | Sensitivity | Specificity | PPV | NPV | n positive | n negative |
|---|---|---|---|---|---|---|---|---|---|
| All Races | Year 1 | 0.833 | 0.264 | 0.798 | 0.604 | 0.057 | 0.99 | 119 | 3973 |
| | Year 2 | 0.731 | 0.263 | 0.801 | 0.426 | 0.079 | 0.972 | 166 | 2686 |
| | Year 3 | 0.685 | 0.32 | 0.803 | 0.346 | 0.111 | 0.945 | 198 | 1954 |
| | Year 4 | 0.686 | 0.335 | 0.804 | 0.329 | 0.124 | 0.934 | 204 | 1720 |
| | Year 5 | 0.687 | 0.409 | 0.802 | 0.344 | 0.171 | 0.912 | 217 | 1287 |
| | Year 6 | 0.685 | 0.46 | 0.796 | 0.367 | 0.25 | 0.871 | 230 | 866 |
| White | Year 1 | 0.963 | 0.453 | 0.786 | 0.957 | 0.355 | 0.993 | 28 | 932 |
| | Year 2 | 0.742 | 0.442 | 0.81 | 0.435 | 0.085 | 0.972 | 42 | 644 |
| | Year 3 | 0.693 | 0.396 | 0.806 | 0.328 | 0.137 | 0.928 | 62 | 470 |
| | Year 4 | 0.692 | 0.409 | 0.806 | 0.333 | 0.15 | 0.922 | 62 | 426 |
| | Year 5 | 0.694 | 0.48 | 0.794 | 0.355 | 0.204 | 0.892 | 68 | 327 |
| | Year 6 | 0.683 | 0.531 | 0.8 | 0.341 | 0.271 | 0.848 | 70 | 229 |
| Black | Year 1 | 0.754 | 0.227 | 0.802 | 0.414 | 0.043 | 0.985 | 91 | 2786 |
| | Year 2 | 0.684 | 0.228 | 0.806 | 0.316 | 0.072 | 0.961 | 124 | 1881 |
| | Year 3 | 0.671 | 0.316 | 0.794 | 0.282 | 0.099 | 0.933 | 136 | 1370 |
| | Year 4 | 0.671 | 0.328 | 0.803 | 0.315 | 0.123 | 0.93 | 142 | 1187 |
| | Year 5 | 0.676 | 0.403 | 0.799 | 0.387 | 0.18 | 0.919 | 149 | 884 |
| | Year 6 | 0.673 | 0.459 | 0.8 | 0.427 | 0.283 | 0.883 | 160 | 567 |

**eFigure 6 Pareto Front of Global ROC-AUC versus Black ROC-AUC performance**

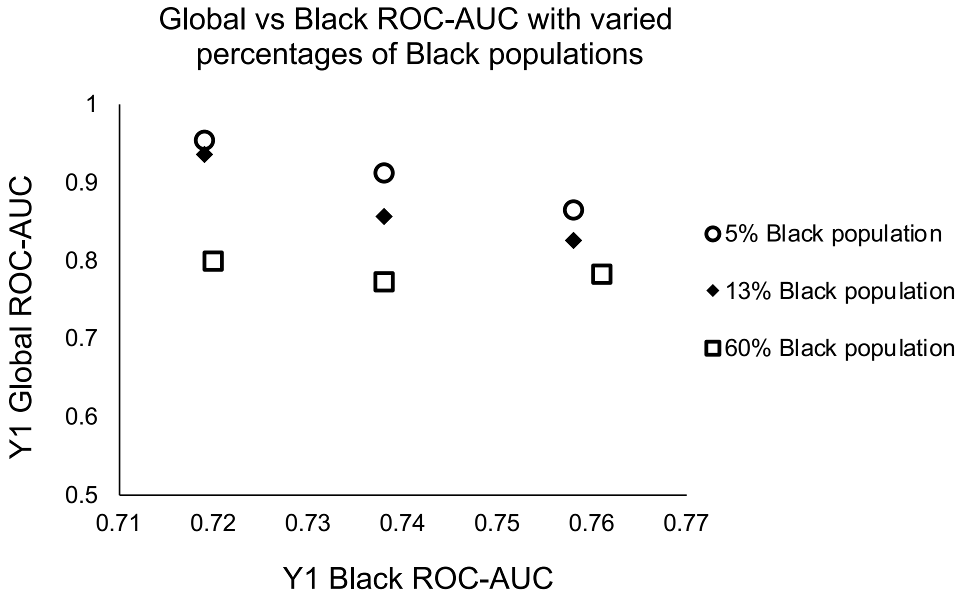

**eFigure 6 –** Pareto Front plot of estimated model Global ROC-AUC performance when optimizing existing ResNet for Black participants (x-axis) in varying populations of 5% (NLST), 13% (US Population), and 60% (UIH Lung cancer screening population).
